# Comprehensive Genetic Study of Russian Hypertrophic Cardiomyopathy Identifies *MYBPC3 c.3697C>T* as the Predominant Variant in Pediatric and Adult Cohorts

**DOI:** 10.1101/2025.11.05.25339569

**Authors:** Leila A. Gandaeva, Olga S. Chumakova, Yulia I. Davidova, Roman P. Myasnikov, Natalia A. Sonicheva-Paterson, Alexander Pushkov, Ilya Zhanin, Daria Chudakova, Alina Alekseeva, Dmitry Demianov, Olga V. Kulikova, Maria M. Kudryavtseva, Daria A. Nefedova, Alina A. Hugaeva, Olga V. Ivleva, Marina I. Berseneva, Alexander Gurshchenkov, Sofia E. Andreeva, Maria Y. Maslova, Natalia S. Krylova, Natalia G. Poteshkina, Dmitry I. Zenchenko, Natalia V. Tverdokhlib, Victoria A. Korneva, Sergey N. Tolstov, Andrey P. Koval, Olga M. Shestopalova, Adilya H. Bukenbaeva, Victoria Khvostikova-Ivanenko, Svetlana E. Glova, Marina V. Gordeeva, Elizaveta A. Yabzhanova, Tatiana A. Romanova, Svetlana N. Nasonova, Anastasia Kruglova, Olga Shchagina, Aleksander Polyakov, Dmitry A. Zateyshchikov, Elena N. Basargina, Sergey Kutsev, Andrey Fisenko, Kirill V. Savostyanov

**Author notes:** **Corresponding author:** Olga Chumakova, Moscow Municipal, Clinical Hospital #17, Volynskaya street, 7, Moscow, 119620, Russia.

## Abstract

**Background.** The genetic etiology of hypertrophic cardiomyopathy (HCM) is complex and population-specific, yet Russian HCM patients remain understudied. **Methods.** Between 2015-2025, unrelated pediatric and adult HCM patients were genotyped in a single laboratory. Children were sequenced with a 404-gene panel and adults with a 17-gene panel. Genotype-phenotype correlations were assessed for carriers of the most frequent variant, *MYBPC3 c.3697C>T*, and compared with patients harboring other *MYPBC3* variants. Haplotype analysis explored its potential founder effect. **Results.** The cohort comprised 315 children (mean age 11.7 years, 61% male; 127 infant-onset) and 3,409 adults (mean age 47.5 years, 58% male). Genotype-positive rates were 89% in children and 28% in adults. Sarcomeric and RASopathy variants accounted for 35% and 33% of infant-onset, 56% and 17% of childhood-onset, and 22% and 0.4% of adult-onset genotyped cases, respectively. The *MYBPC3 c.3697C>T* variant was identified in 83 patients (2.2%). Compared with controls (n = 85), carriers (n = 58) more often had left ventricular ejection fraction below 50% (9% vs. 1%, p = 0.038), left atrial dilation (76% vs. 47%, p = 0.001), and supraventricular arrythmias (45% vs. 25%, p = 0.027). Pediatric carriers showed no left ventricular outflow tract obstruction (0% vs. 29%, p = 0.024) and fewer conduction disturbances (13% vs. 48%, p = 0.029). Intervention rates and outcomes were similar across ages. Haplotype analysis was inconclusive for founder effect. **Conclusions.** The genetic spectrum of HCM in Russian population largely mirrors Western data. Within *MYBPC3*-associated HCM, the *c.3697C>T* variant is linked to a milder, nonobstructive pediatric phenotype and increased hypokinetic progression in adults but does not influence overall outcomes. Its high prevalence likely reflects a combination of founder and recurrent mutational events.

## Introduction

Hypertrophic cardiomyopathy (HCM) is the most common hereditary cardiac disorder, with a prevalence of 1:200 to 1:500 (1,2). It is defined by left ventricular (LV) hypertrophy not explained solely by abnormal loading conditions (3) and demonstrates substantial clinical heterogeneity. Although the median age at diagnosis is approximately 45 years (4), 5-15% of patients present during childhood (5,6), yet pediatric data remain scarce. Clinical spectrum ranges from asymptomatic to severe manifestations, including progressive heart failure (HF) or sudden cardiac death (SCD) predominantly in individuals <35 years. This variability likely results from interplays between primary HCM-associated nucleotide variants, environmental, and non-Mendelian genetic factors (7).

Pathogenic (P) and likely pathogenic (LP) variants in the sarcomeric genes account for 30-60% of HCM cases, most commonly myosin binding protein C (*MYBPC3*) and myosin heavy chain 7 (*MYH7*).

Non-sarcomeric HCM shows some differences between adults and children. Adults are often genotype-elusive; only minority carry variants in non-syndromic sarcomere-associated genes (around 10%) or syndromic genes (around 5%), mainly alpha-galactosidase A (*GLA*) and transthyretin (*TTR*), responsible for Fabry disease and hereditary amyloidosis. In contrast, pediatric HCM shows higher prevalence and variability of syndromic phenocopies (15-40% depending on age of onset), including RASopathies, inborn errors of metabolism (IEM), and neuromuscular diseases (8,9).

Most genetic data in cardiomyopathies derive from Western populations of European ancestry. Meanwhile, underrepresented populations may exhibit distinct variant frequency profiles, thereby complicating pathogenicity assessment and clinical decision-making. Recurrent variants, often originating from common ancestors (the ‘founder effect’), are of particular interest as incomplete penetrance allows them to persist through generations. Cataloging such variants across populations would enable more accurate estimates of their population-level penetrance for considering how such incidental findings should be reported and followed in future whole-genome sequencing (10). In HCM, *MYBPC3* is the gene most frequently reported with founder variants.

To date Russian HCM patients mostly remain understudied. Only a few cohorts have been reported, including 68 pediatric cases limited to infantile onset (diagnosed within the first year of life) – a subgroup substantially distinct from most HCM presentations (9) – and 176 adult patients (11), in whom *MYBPC3 c.3697C>T* (*p.Q1233\**) was the most common variant (4.6%), exceeding *MYBPC3 c.1504C>T* (*p.R502W*), which predominates in Western populations (12). The *c.3697C>T* variant was associated with earlier disease onset (11), but this investigation was conducted at a single center with a limited sample size, and it is still uncertain whether *c.3697C>T* constitutes a founder mutation within the Russian population.

In this study, we recruited the largest cohort of Russian HCM patients to (i) characterize the genetic spectrum in pediatric and adult groups and determine the prevalence of the *MYBPC3 c.3697C>T* variant, (ii) assess genotype-phenotype correlations among its carriers, and (iii) explore a potential founder effect.

## Material and Methods

### Pediatric cohort

Between Jan 2015 and Jan 2025, 1007 unrelated children (aged ≤ 17 years) with various cardiomyopathies were followed and genotyped at the Laboratory of Medical Genomics National Medical Research Center for Children’s Health (Moscow, Russia). Of these, 315 children with an HCM phenotype were included. HCM was established according to current guideline criteria for LV wall thickness, adjusted for body size and growth (3). Final diagnoses, integrating clinical and genetic data, were available for all pediatric patients.

### Adult cohort

Between Nov 2020 and Jan 2025, 3,409 seemingly unrelated patients with HCM (aged ≥ 18 years) were genotyped at the same laboratory, National Medical Research Center for Children’s Health, where the pediatric cohort was studied. The initial HCM diagnosis was provided by referring physicians from over 20 centers across Russia. Final diagnostic data were unavailable for most adults.

### Genetic study design

All children underwent genetic testing using a custom-built, previously described targeted next-generation sequencing panel of 404 genes implicated in hereditary cardiovascular diseases (13) (**Figure 1A**). The target regions comprised both coding sequences and the adjacent intronic regions. Based on the most frequent findings in children older than 1 year of age, a streamlined 17-gene panel was developed for HCM adults, primarily to screen for Fabry disease (**Supplementary Table 1**). Technical details of the genetic analyses are provided in the **Supplementary File**. The *MYBPC3 c.3697C>T* variant was confirmed in all carriers by Sanger sequencing. Variants were classified per 2015 American College of Medical Genetics and Genomics (ACMG) criteria (14). Patients carrying P/LP variants or variants of uncertain significance (VUSs) with high pathogenic potential were considered genotype-positive.

**Figure 1.**
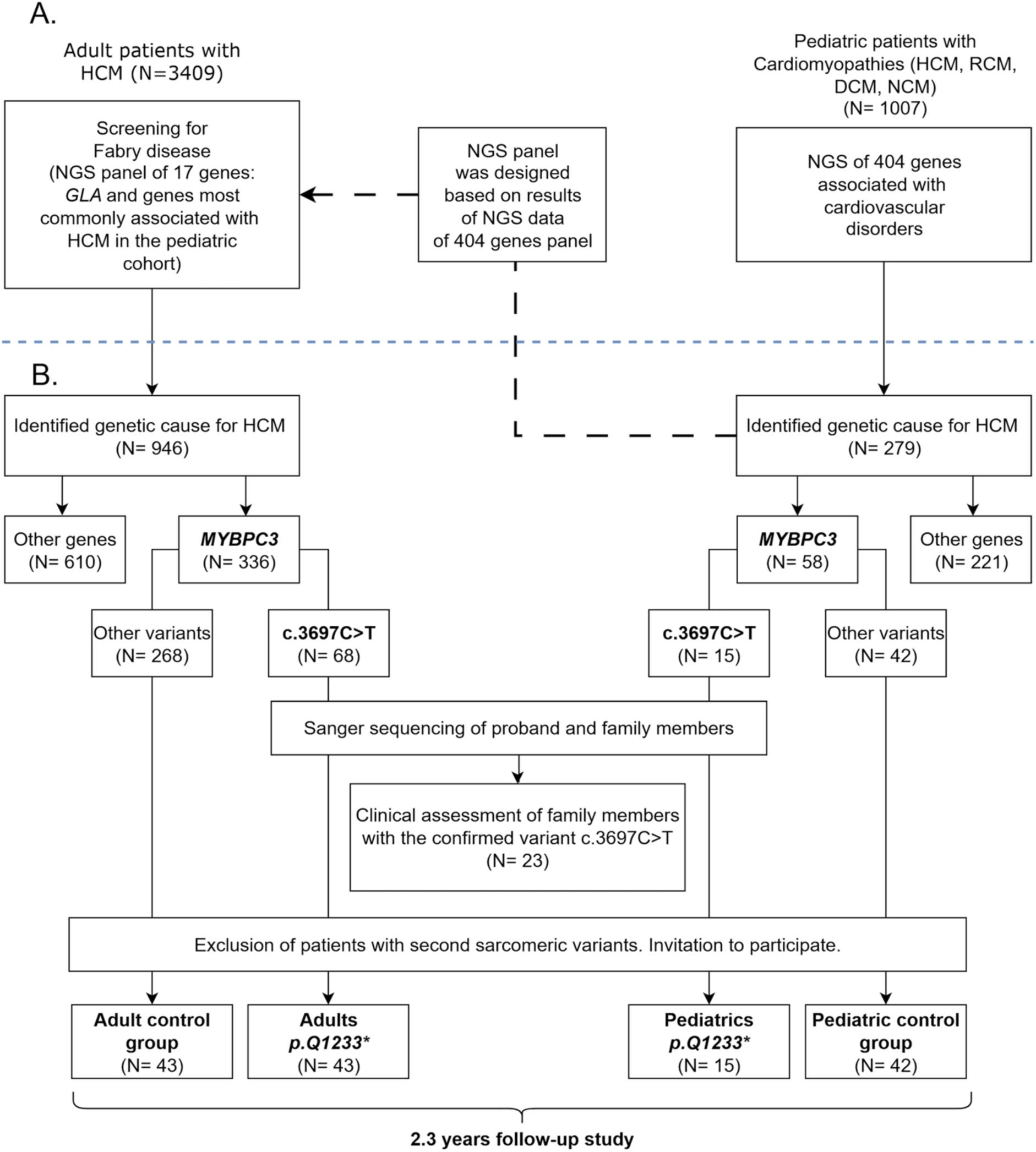
Flowchart. (A) Genetic analysis of pediatric and adult Russian HCM patients. (B) Design of the genotype-phenotype study for the *MYBPC3 c.3697C>T* variant. NGS - next-generation sequencing, HCM – hypertrophic cardiomyopathy, RCM – restrictive cardiomyopathy, DCM – dilated cardiomyopathy, NCM – non-compaction myocardium.

### MYBPC3 c.3697C>T cohort: genotype-phenotype correlation analysis

Pediatric and adult patients with HCM who carried the *MYBPC3 c.3697C>T* variant were selected for genotype-phenotype analysis (**Figure 1B**). Patients with additional P/LP sarcomeric variants were excluded. Clinical data for pediatric patients were obtained from the National Medical Research Center for Children’s Health records, and data for adults were collected through invitations to referring clinicians (**Supplementary Table 2**).

The control group comprised HCM patients carrying a single P/LP variant or a VUS with high pathogenic potential in *MYBPC3* (**Supplementary Table 3**). Pediatric controls were recruited at the National Medical Research Center for Children’s Health, whereas adult controls at Moscow Municipal Clinical Hospital #17 and the National Medical Research Center for Therapy and Preventive Medicine.

Family screening was performed whenever possible, and relatives carrying the *MYBPC3 c.3697C>T* variant who met HCM diagnostic criteria were invited to participate (**Figure 1B**).

Comparative analyses of baseline characteristics, intervention rates, and clinical outcomes were conducted between the *MYBPC3 c.3697C>T* cohort and controls, stratified by age. Data were entered into a study-specific electronic case report form. Baseline clinical information included demographics, personal and family history, electrocardiographic findings, and cardiac imaging parameters. Outcomes, collected retrospectively, comprised all-cause mortality, SCD, heart transplantation, worsening HF, new-onset stroke, and a composite outcome encompassing all of these events. Worsening HF was defined as hospitalization requiring parenteral diuretics and/or inotropes, or progression to New York Heart Association (NYHA) class III/IV. Interventions, including surgical myectomy and implantation of cardioverter-defibrillators or pacemakers, were documented at baseline and during follow-up.

### MYBPC3 c.3697C>T cohort: haplotype analysis

Haplotype analysis was performed on 48 patients with *MYBPC3 c.3697C>T* variant and 50 unrelated participants from the Russian population (control group). The allele frequencies of the *c.3697C>T* (coordinate 47,354 Kb) and of four microsatellite markers in both homozygous and heterozygous states were determined. Microsatellite markers included D11S4174 (coordinate 45,282Kb), D11S1344 (coordinate 46,167Kb), D11S4109 (coordinate 47,601Kb), and D11S1978 (coordinate 48,593Kb), which flank the *MYBPC3* gene at its the 5’ and 3’ ends. All coordinates are given based on genomic assembly hg37/19. All studied markers are located within the same 58.40 cM region according to the Marshfield Comprehensive human genetic maps.

The study was approved by the Local Ethics Committee of the National Medical Research Center for Children’s Health. Written informed consent for the use of medical records in research and publication was obtained from pediatric patients’ legal guardians or next of kin, and from all adult participants through their treating clinicians.

#### Statistical analysis

Categorical variables were expressed as counts and percentages and analyzed using Pearson’s χ² test or Fisher’s exact test (for expected frequencies <5). Continuous variables were expressed as mean ±SD or as median and interquartile range [IQR, 25th-75th percentiles] and compared using Student’s t-test (for normally distributed data, assessed by Shapiro-Wilk test) or Mann-Whitney U-test. For haplotype analysis, **Х^2^** test was used for pairwise comparisons of allele frequencies between patient and control samples; to compare alleles in two samples, **Х^2^** test was used with the number of degrees of freedom corresponding to the number of alleles of particular marker minus 1. Following Rothman’s recommendations (15), adjustments for multiple testing were not applied, to avoid obscuring potentially important biological relationships given the limited sample size. Analyses were performed using SPSS Statistics v28 (IBM Corporation), with two-tailed p-values < 0.05 considered significant.

## Results

### Pediatric HCM cohort

Of 315 children with HCM (mean age 11.7 ± 4.3 years; mean age at diagnosis 5.0 ± 5.7 years; 61% male), 125 (40%) had infant-onset and 190 (60%) childhood-onset disease. Overall, 87 (28%) presented with LV outflow tract obstruction (LVOTO), and 90 (29%) had a familial history of cardiomyopathy.

A total of 230 unique variants were identified in 279 children, corresponding to a genotype-positive rate of 88.6%. Ten patients (4%) harbored multiple variants: eight carried double sarcomeric variants, while two had *MYH7*+*FLNC* and *MYH7*+*PRKAG2* combinations. *MYH7* variants were the most frequent (82/290, 28%), followed by *MYBPC3* (62/290, 21%). Separate analyses were performed for infant- and childhood-onset groups (**Figure 2**).

**Figure 2.**
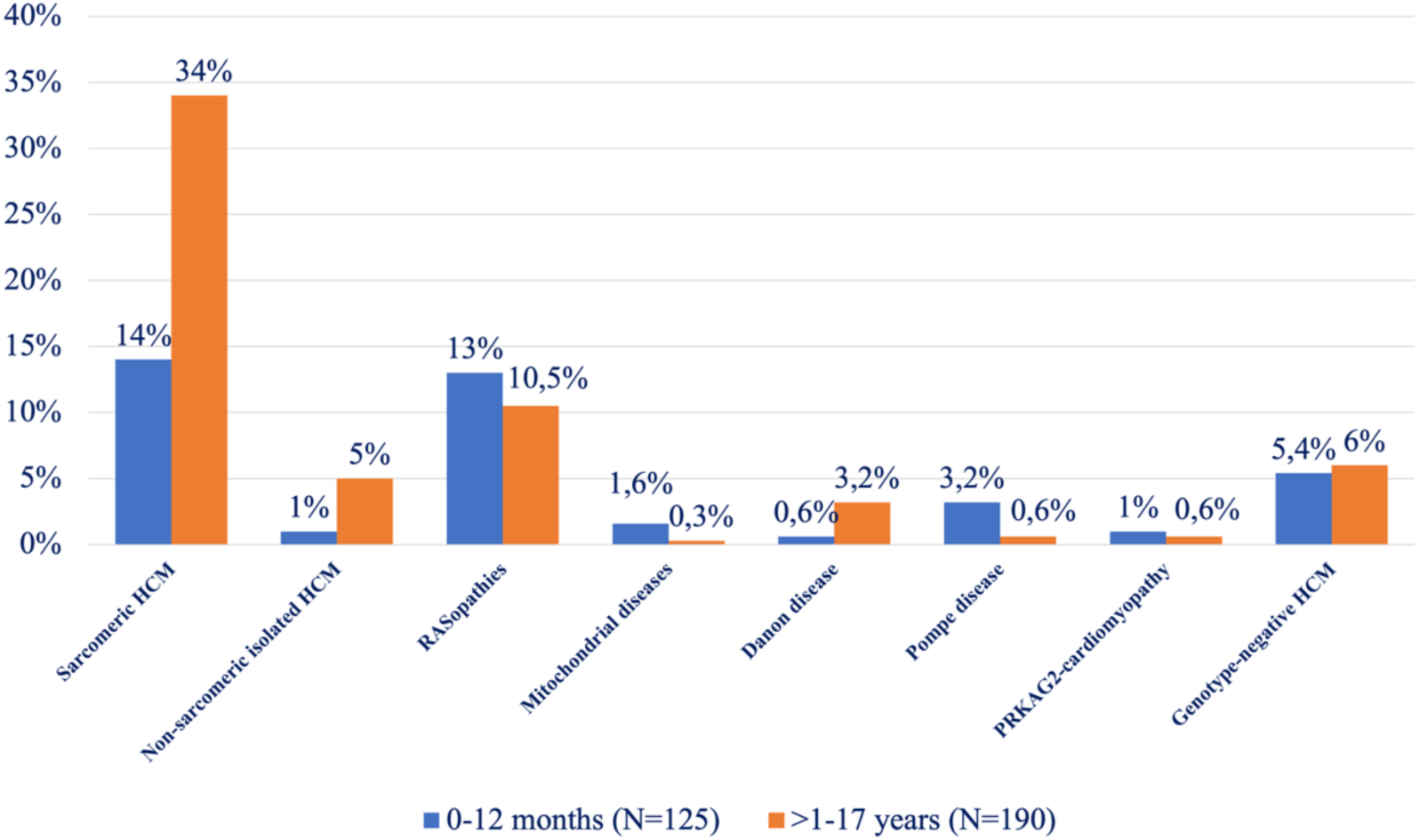
Distribution of final diagnoses in Russian pediatric HCM patients (N = 315) with respect to age of onset. The percentages indicate the proportion of diagnoses within the total group.

Sarcomeric HCM was more frequent in childhood-onset than in infant-onset patients (107/190, 56% vs. 44/125, 35%, p < 0.0001). The prevalence of non-sarcomeric isolated HCM was similar between groups (35/190, 18% vs. 20/125, 16%, p=0.7); however, most of these infant-onset cases remained genotype-negative (17/20, 85%), whereas almost half of the childhood-onset cases (16/35, 46%) had identifiable variants. Variants occurred across several gene groups (numbers in parentheses denote carriers): cytoskeletal/sarcomere-related (*DES* (1)*, FHL1* (1)*, FHOD3* (2)*, FLNC* (4), *JPH2* (2)*, TCAP* (1)*, TRIM63* (1)*, TTN* (1)), ion-channel (*CACNA1C* (2), *RYR2* (1), *SCN5A* (1)), and signaling/regulatory (*AKAP9* (1), *PRDM16* (1)). All four *FLNC* carriers (three missense, one in-frame deletion) were childhood-onset with benign courses, without restrictive physiology, myocardial fibrosis, or arrythmias. The deletion carrier showed marked LV hypertrophy (29 mm) and elevated N-terminal pro-brain natriuretic peptide (1143 pg/mL).

Syndromic HCM was twice as prevalent in infant-onset compared to childhood-onset patients (61/125, 49% vs. 48/190, 25%, p < 0.0001). RASopathies and mitochondrial disorders were more frequent in infant-onset HCM: 41/125, 33% vs. 33/190, 17% (p = 0.002) and 5/125, 4.0% vs. 1/190, 0.5% (p = 0.07), respectively. RASopathy variants were identified in *BRAF* (6)*, HRAS* (2), *KRAS* (1)*, LZTR1* (4)*, NF1* (1), *PTPN11* (23), *RAF1* (19), *RIT1* (10)*, SHOC2* (2), *SOS1* (6); mitochondrial variants in *ACAD9* (3)*, ELAC2* (1)*, MTO1* (1)*, POLG1* (1). Among metabolic/storage disorders, Pompe disease and *PRKAG2*-cardiomyopathy predominated in infant-onset HCM (10/125, 8% vs. 2/190, 1.1%, p = 0.004 and 3/125, 2.4% vs. 2/190, 1.1%, p = 0.64), while Danon disease was more frequent in childhood-onset HCM (10/190, 5.3% vs. 2/125, 1.6%, p = 0.17). No pediatric patients had *GLA* variants associated with Fabry disease.

### Adult HCM cohort

Of 3,409 adult HCM patients (mean age 47.5 ± 0.4 years; range 18.6 – 84.8 years; 58% male), 946 (28%) carried at least one rare variant in 17 HCM-associated genes. Multiple variants, including at least one sarcomeric, were found in 49/946 patients (5.2%), and one had two variants in non-sarcomeric *GLA* and *LAMP2* genes. **Figure 3** shows the distribution of variants by gene. Sarcomeric variants accounted for 78% of findings (777/996); overall, 749/3,409 patients (22%) carried sarcomeric variants. *MYBPC3* was most frequent (336/996, 34%) followed by *MYH7* (293/996, 29%). *FLNC* variants occurred in 114/3,409 patients (3.3%), all missense; 11 carriers (10%) also had a sarcomeric variant. In 18/3,409 patients (0.5%), *FLNC* variants were classified as P/LP. *DES* variants (16 missense, one nonsense, and one splicing) were found in 27/3,409 patients (0.8%), and in 11/3,409 cases (0.3%) were classified as P/LP (**Supplementary Table 4**); four carried additional sarcomeric variants. Variants in genes related to hereditary amyloidosis (*TTR,* n=19), Fabry disease (*GLA,* n=11), *PRKAG2*-cardiomyopathy (*PRKAG2,* n=17), Danon disease (*LAMP2,* n=14), and Noonan syndrome (*PTPN11,* n=14) were rare, collectively identified in 74 patients, representing 8% of genotype-positive patients and 2% of the total genotyped HCM cohort.

**Figure 3.**
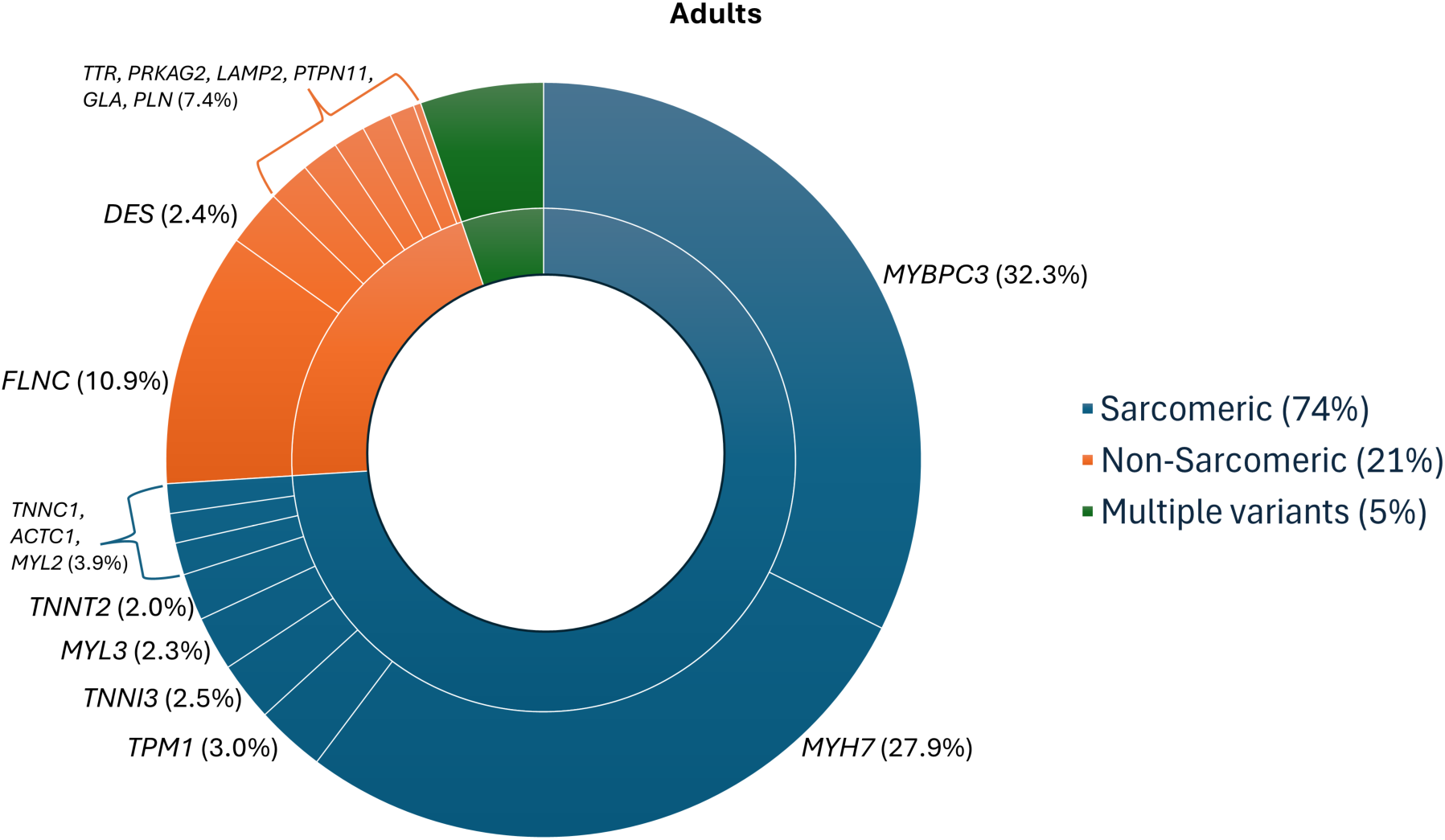
Distribution of identified variants by gene in Russian adult HCM patients (N = 946). Single sarcomeric variants are shown in navy blue, single non-sarcomeric variants in orange, and multiple variants in green.

### MYBPC3 c.3697C>T cohort: genotype-phenotype correlations

Among 3,702 unrelated pediatric and adult HCM patients, the *MYBPC3 c.3697C>T* variant was present in 2.2% (83 cases): 15/315 children (5.0%) and 68/3,409 adults (2.0%). Family screening was conducted for 28 relatives across 16 families, resulting in the identification of 23 additional carriers assessed for HCM phenotype. The final *c.3697C>T* study group included 58 eligible patients without second sarcomeric variants whose clinicians consented to participate: 15 children and 43 adults, including 38 probands and 5 relatives (**Figure 1B**). Probands represented 91% of the group; the only significant difference between adult probands and relatives was younger age at diagnosis in the latter (31±6 vs. 41±16 years, p = 0.026). The control group included 85 patients with other *MYBPC3* variants: 42 children and 43 adults (37 probands, 6 relatives). Baseline clinical characteristics are in **Supplementary Table 5.**

#### Adult vs. pediatric c.3697C>T -associated HCM

Compared with adults (median age 38 years), pediatric carriers (median age 11 years, three diagnosed in infancy) were more often male (73 vs. 37%, p = 0.019), had milder LV hypertrophy (17±5 vs. 22±5 mm), higher LV ejection fraction (74 vs. 66%, p < 0.0001), and none showed LVOTO (0 vs. 38%, p = 0.006) or elevated pulmonary artery pressure (0 vs. 39%, p = 0,005). Adults more frequently had paroxysmal supraventricular tachycardia (61 vs. 0%, p < 0.0001), myocardial fibrosis on cardiac magnetic resonance imaging (95 vs. 33%, p = 0.005), and more often required diuretics (44 vs. 0%, p = 0.001). Non-significant trends (p = 0.06) included a higher rate of Wolff-Parkinson-White pattern on electrocardiogram in children (13 vs. 0%) and non-sustain ventricular tachycardia in adults (37 vs. 7%).

#### Study group vs. controls

Compared with controls, the *c.3697C>T* carriers more often exhibited a hypokinetic disease stage (9 vs. 1%, p = 0.038), left atrial dilation (76 vs. 47%, p = 0.001), and supraventricular arrythmias (45 vs. 25%, p = 0.027). Comparison between adult subgroups showed no significant differences. Compared with pediatric controls, children with the *c.3697C>T* had no LVOTO (0 vs. 29%, p = 0.024), fewer conduction disturbances (13 vs. 48%, p = 0.029), but more frequent left atrial dilation (60 vs. 21%, p = 0.014). Intervention rates or outcomes did not differ between pediatric and adult carriers or between the study group and controls (**Supplementary Table 6**).

### Haplotype analysis

Out of 48 patients, reliable results were achieved for 46, with pre-analytical issues affecting two samples. Due to the incomplete availability of biological samples from the patient’s family members, accurate determination of the haplotypes associated with *c.3697C>T* variant was not feasible. Alleles “4” “10” “9” and “3” for markers D11S4174, D11S1344, D11S4109, D11S1978, respectively, were found with the highest frequency among patients with heterozygous *c.3697C>T* variant, but none of the alleles for any of the markers was detected in all examined patients (**Table 1**). The allele frequencies of microsatellite markers of 0.5 and higher were not due to the presence of allele in all patients, but because of its presence in the homozygous state in some patients.

**Table 1.** Allele frequencies of microsatellite markers in patients with *c.3697C>T* variant in *MYBPC3* and controls. The allele frequencies of the most frequent alleles are highlighted in gray.

| Allele | D11S4174 |  | D11S1344 |  | D11S4109 |  | D11S1978 |  |
| --- | --- | --- | --- | --- | --- | --- | --- | --- |
|  | patients | controls | patients | controls | patients | controls | patients | controls |
| 1 | 0.01 | 0 | 0.01 | 0.00 | 0.00 | 0.00 | 0.01 | 0.00 |
| 2 | 0.05 | 0.04 | 0.03 | 0.00 | 0.00 | 0.02 | 0.06 | 0.15 |
| 3 | 0.33 | 0.24 | 0.14 | 0.07 | 0.12 | 0.10 | 0.57 | 0.35 |
| 4 | 0.50 | 0.4 | 0.01 | 0.02 | 0.00 | 0.04 | 0.18 | 0.25 |
| 5 | 0.05 | 0.06 | 0.03 | 0.09 | 0.08 | 0.06 | 0.04 | 0.02 |
| 6 | 0.07 | 0.2 | 0.13 | 0.25 | 0.01 | 0.08 | 0.02 | 0.02 |
| 7 | 0.00 | 0.02 | 0.03 | 0.05 | 0.08 | 0.10 | 0.00 | 0.00 |
| 8 | 0.00 | 0.04 | 0.15 | 0.36 | 0.06 | 0.02 | 0.05 | 0.10 |
| 9 | 0.00 |  | 0.08 | 0.00 | 0.47 | 0.37 | 0.00 | 0.06 |
| 10 | 0.00 |  | 0.32 | 0.16 | 0.03 | 0.10 | 0.01 | 0.00 |
| 11 | 0.00 |  | 0.06 | 0.00 | 0.02 | 0.04 | 0.05 | 0.00 |
| 12 | 0.00 |  | 0.01 | 0.00 | 0.13 | 0.06 | 0.05 | 0.04 |
| 13 | 0.00 |  | 0.00 | 0.00 | 0.00 | 0.04 | 0.00 | 0.00 |

Among the 46 patients examined in our *MYBPC3 c.3697C>T* cohort, all but one also carried the *MYBPC3 c.977A* variant. The patients’ genotype was as follows: D11S4174-D11S1344-c.977G>A-c.3697 C>T-D11S4109-D11S4109-D11S4109: 4/4 (50%)-1/10 (1%/32%)-G/G (1%)-T/C (50%/50%)-9/9(47%)-3/3 (57%) (brackets indicate allele frequency in % in patients with the *c.3697C>T* variant). Allele frequencies of *MYBPC3 c.3697C>T* and *c.977G>A* variants were determined in our patients and in the Database of Russian population allele frequencies (RuExac) (**Table 2**).

**Table 2.**
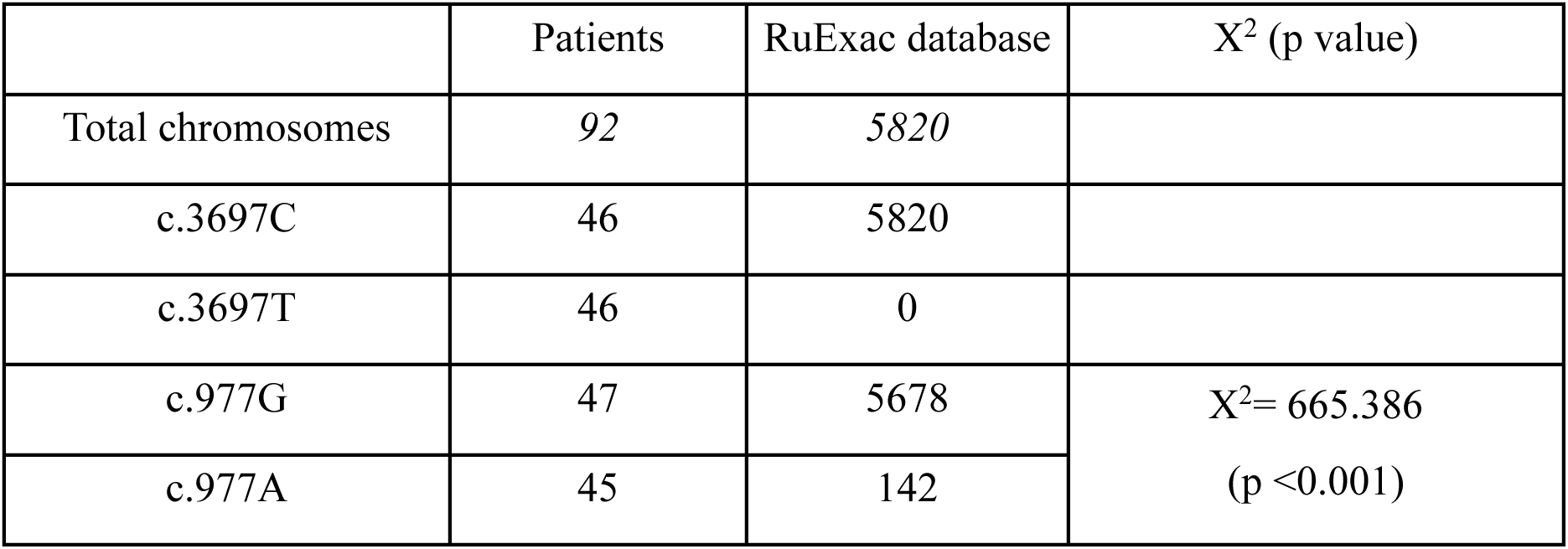
Allele frequencies of *c.3697C>T* and *c.977G>A* variants in *MYBPC3* in our patients and RuExac database.

## Discussion

### Genetic findings in Russian HCM patients

We present the most comprehensive data to date on the genetic spectrum of HCM in the Russian population, encompassing a large pediatric cohort. Consistent with previous studies, genetic testing yield was higher in children than in adults. *MYBPC3* and *MYH7* were the main genes involved, with *MYH7* variants more frequent in children and *MYBPC3* predominating in adults (6,16), reflecting the typically later onset of *MYBPC3*-associated disease.

The high genotype-positive rate observed in our pediatric groups (86-90%) is consistent with recent reports (6,9,17), but exceeds the largest comparable studies by Norrish and Kaski et al. (60-70%) (8,18). This may result from the use of a single laboratory and an expanded gene panel in the present study. In contrast, the modest genetic yield in adults (28%) likely relates to the unrestricted inclusion of older patients and a more limited gene panel.

As in other populations (6,18) and a separate Russian cohort (9), our infant-onset patients were diagnosed with sarcomeric HCM in one-third cases – a substantial proportion, making disease etiology in this group closer to that in adults (19). In total, non-syndromic HCM was present in half of our youngest patients, slightly higher than previous reports (35-45%) (17,18). The proportion of RASopathies is stable across studies (30-35%; 33% in our group) (9,17,18), while IEM accounted for 16%, consistent with Norrish et al. (18) but within a broader 10-27% range in other studies (9,17).

In our childhood-onset HCM, sarcomeric forms were 1.5 times more frequent than in infants (56%), similar to Marston et al. (63%) (6) and higher than in Kaski et al. (40%) (8). Compared with infants, the prevalence of newly diagnosed RASopathies and IEMs decreased twice with age (33% vs. 17% and 16% vs. 8%), matching rates in study by Kaski et al. (18% and 8%) for children aged 1-18 years (8).

All Fabry disease cases were diagnosed in adults, consistent with evidence that cardiac involvement in non-classic Fabry disease manifests later in life (20).

*FLNC* variants were found in 3.3% of adults and 2.1% of childhood-onset HCM patients, consistent with prior reports (8,21). We did not observe *FLNC* variants in infant-onset patients, as was noted by Fetisova et al. (9). *FLNC* contributes to HCM mainly through non-loss-of-function missense variants (22), and our findings support this. Some variants were classified as P/LP (0.5% of adults), but lacking segregation and functional data precluded reliable estimation of P/LP *FLNC* variant frequency in Russian HCM. Although genotype-phenotype correlations were beyond this study’s scope, follow-up of four children and two previously described adults (11) suggested a benign course, consistent with previous observations (21).

The *DES* gene has only recently been validated for inclusion in HCM panels by the ClinGen Hereditary Cardiovascular Disease Gene Curation Expert Panel (22). Among cardiomyopathies, HCM is the rarest cardiac phenotype associated with *DES* variants. Consequently, data on *DES*-related HCM remain scare. Although clinical information was unavailable for most adult patients in our cohort, reporting *DES* variants, including novel ones, will contribute to expending knowledge in this field.

### Genotype-phenotype study of the MYBPC3 c.3697C>T variant

To our knowledge, this study investigated the largest cohort of HCM patients carrying the *MYBPC3 c.3697C>T* variant, the most common in the Russian HCM population (prevalence 2.2%). By contrast, in Western countries, its prevalence is reported at less than 1% (4,16).

In our cohort, *c.3697C>T*-associated HCM did not differ from other *MYBPC3*-associated HCM in major outcomes, and age of onset did not affect prognosis. Compared to adults, children more often carried this variant (one-third of MYBPC3-associated cases), were predominantly male, and showed a milder phenotype – less LV hypertrophy, higher LV ejection fraction, and no LVOTO. These findings suggest age-dependent expression with limited early-life impact, indicating that pediatric carriers may require long-term follow-up rather than immediate intervention. The high rate of hypokinetic stage (9%) and myocardial fibrosis (95%) in adults may warrant closer HF monitoring, though these findings need further confirmation.

Comparison with the only previously characterized Belarusian cohort (23) revealed no major differences. Future studies in larger Slavic cohorts may provide more robust data.

### Haplotype analysis of MYBPC3 c.3697C>T variant

The *c.3697C>T* variant in *MYBPC3* has been reported as a founder mutation in the Belarusian population ethnically related to Russia (23). Two additional reports from non-Slavic groups provided no convincing evidence: Erdmann et al. identified it as a founder variant among individuals of Turkish descent based on haplotype analysis of two unrelated patients (24), while Sepp et al. reported 12 Hungarian HCM cases suggesting a founder effect, though without haplotype confirmation (25).

In our study, 45 of 46 patients with *c.3697C>T* also carried *c.977G>A*, a co-occurrence previously reported in Russian and Belarusian cases (23,26). Family studies confirmed the cis-position of these variants (23,27). Further, while the *c.3697C>T* is absent in the RuExac, the *c.977G>A* appears with an allele frequency of 2.44% (two homozygotes).

A significant difference in the *c.977A* allele frequency between patient’s chromosomes carrying and lacking the *c.3697C>T* (p<0.001) may indicate the founder effect, suggesting that *c.3697C>T* likely arose on a chromosome already containing the *c.977A* and subsequently expanded in Slavic populations. Erdmann et al. described a similar founder haplotype in Turkish patients using microsatellite markers D11S1385 and D11S1313, although the status of *c.977G>A* variant was unreported (24).

In our study, the allele frequencies for markers D11S4174 (allele 4), D11S4109 (allele 9), and D11S1978 (allele 3) were similar between patients and controls. This lack of difference may reflect that disease-causative *c.3697C>T* variant occurs on one of the most common Russian haplotypes. However, the presence of different alleles of the same markers among patients with *c.3697C>T* variant suggests an ancient mutational event. Multiple independent mutational events cannot be excluded as well; this would be confirmed if many patients carried the *c.3697T* but without the *c.977A*. Notably, one of 46 patients did not have *c.977A* variant but carried microsatellite marker alleles which were common in other patients and the general population.

Overall, our data neither confirm nor exclude a founder effect for *c.3697C>T* variant in *MYBPC3* gene in Russian HCM patients. The findings most likely indicate a combination of founder effect and recurrent mutational events contributing to its accumulation in this population.

#### Conclusions

The genetic spectrum of HCM in Russian population largely mirrors Western data. The use of expanded genetic panel as a first-line approach in pediatric HCM yielded a high rate of positive results. In the rare infant-onset subgroup, sarcomeric and RASopathy variants accounted for 70% of cases in equal proportions. In *MYBPC3*-associated HCM, the *c.3697C>T* variant was linked to a milder, nonobstructive pediatric phenotype and increased hypokinetic progression in adults, without affecting overall outcomes. Our findings suggest that both founder effect and recurrent mutational events contributed to *c.3697C>T* accumulation in Russian HCM population. This cohort represents a valuable, genetically homogenous resource for studying phenotype modifiers in HCM.

#### Limitations

Adult patients were tested with a limited gene panel, so some non-sarcomeric variants may have been underestimated; clinical data were unavailable for most adults, restricting accurate variant pathogenicity classification; the retrospective genotype-phenotype analysis did not allow assessment of *c.3697C>T* variant penetrance.

## Supporting information

Supplemental Tables 1-6 and Technical details of the genetic study

## Data Availability

All data produced in the present study are available upon reasonable request to the authors

## Acknowledgments

None

## Sources of Funding

None

## Disclosures

None

## Notes

### Competing Interest Statement

The authors have declared no competing interest.

### Funding Statement

This study did not receive any funding

### Author Declarations

Ethics committee/IRB of National Medical Research Center for Children Health gave ethical approval for this work.

