## Supplemental Tables 1-6 and Technical details of the genetic study for "Comprehensive Genetic Study of Russian Hypertrophic Cardiomyopathy Identifies *MYBPC3 c.3697C>T* as the Predominant Variant in Pediatric and Adult Cohorts"

**Supplementary Table 1.** Custom-built 17-gene panel for hypertrophic cardiomyopathy in adults

| *ACTC1* | Actin, alpha cardiac muscle 1 |
| --- | --- |
| *DES* | Desmin |
| *FLNC* | Filamin-C |
| *GLA* | Alpha galactosidase A |
| *LAMP2* | Laminin subunit alpha-2 |
| *MYBPC3* | Myosin-binding protein C, cardiac-type |
| *MYH7* | Myosin Heavy Chain 7 |
| *MYL2* | Myosin regulatory light chain 2, ventricular/cardiac muscle isoform |
| *MYL3* | Myosin light chain 3 |
| *PLN* | Cardiac phospholamban |
| *PRKAG2* | 5'-AMP-activated protein kinase subunit gamma-2 |
| *PTPN11* | Tyrosine-protein phosphatase non-receptor type 11 |
| *TNNC1* | Troponin C1, Slow Skeletal and Cardiac Type |
| *TNNI3* | Troponin I3, Cardiac Type |
| *TNNT2* | Troponin T2, Cardiac Type |
| *TPM1* | Tropomyosin 1 |
| *TTR* | Transthyretin |

**Technical details of the genetic study**

Genomic DNA was isolated from 200 ul of peripheral blood using DNA Blood Mini Kit (QIAGEN, Germany) on automated DNA isolation station QIAQUBE (QIAGEN, Germany) and from dried blood spots using MagPure Universal DNA Kit (Magen, China) on automated DNA isolation station Auto-Pure 96 (Allsheng, China). The quality and quantity of DNA was assessed via spectrophotometry using NanoPhotometer N60 (Implen, Germany) or Qubit dsDNA HS Assay Kit and Qubit 3.0 fluorometer (Invitrogen, USA).

Libraries for next-generation sequencing were prepared using a KAPA HyperPlus Kit (Roche, USA) according to the manufacturer’s protocol. The DNA fragmentation time was 15 minutes to achieve an average fragment length of 350 bp. Target enrichment was carried out using KAPA HyperCap hybridization probes (Roche, USA). Massive parallel sequencing was performed on the MiSeq platform (Illumina, USA) with V2 chemistry (500 cycles, paired-end reads) and NextSeq 2000 platform (Illumina, USA) with P3 chemistry (300 cycles, paired-end reads).

Bioinformatic analysis was carried out according to the guidelines of Genome Analysis Toolkit Best Practices (https://gatk.broadinstitute.org). Briefly, raw reads were trimmed using Trimmomatic (version 0.39). Then, sequence alignment was performed with BWA-MEM2 (version 2.2) using GRCh38 genome assembly as a reference. Next, duplicate reads were marked using Picard tools and base quality score recalibration (BQSR) was performed. Then, genetic variations (SNPs and indels) were called with HaplotypeCaller using GATK (version 4.5). Next, gene annotation was performed with in-house script to annotate variations present inClinVar, OMIM and HGMD [<http://www.hgmd.cf.ac.uk>] databases. The pathogenicity of variants not previously described was determined using the Alamut program with the built-in software modules SIFT, PolyPhen HDIV, PolyPhen HVAR, Mutation Taster, FATHMM, CADD13, DANN, M-CAP, REVEL, as well as using the American College of Medical Genetics and Genomics (ACMG) manual. Finally, a filtering process removed variations outside targeted sequences, with a population frequency > 0.5% (gnomAD v3.1.1). The clinical interpretation of the genetic results was done according to a joint consensus recommendation of the ACMG and the Association for Molecular Pathology (AMP) (<https://linkinghub.elsevier.com/retrieve/pii/S1098360021030318>).

Sanger sequencing was performed using the BigDye® Terminator v3.1 Cycle Sequencing Kit (Thermo Fisher Scientific, USA) in accordance with the manufacturer's protocols and guidelines. Amplification was performed on Bio-Rad T100 (Bio-Rad, USA) and ProFlex (Thermo Fisher Scientific, USA) thermocyclers. Capillary electrophoresis was performed on ABI 3500XL automated DNA sequencer (Thermo Fisher Scientific, USA). The obtained sequences were compared with RefSeqGene reference sequences from the National Center for Biotechnology Information database.

**Supplementary Table 2.** List of Centers Participating in the *MYBPC3 c.3697C>T* Variant Genotype-Phenotype Study

National Medical Research Center for Children’s Health, Moscow, Russia

Moscow Healthcare Department, Municipal Clinical Hospital #17, Moscow, Russia

National Medical Research Center for Therapy and Preventive Medicine, Moscow, Russia

Bakoulev Scientific Center for Cardiovascular Surgery, Moscow, Russia

Almazov National Medical Research Centre, Saint-Petersburg, Russia

Pirogov Russian National Research Medical University, Moscow, Russia

Volgograd State Medical University, Volgograd, Russia

Orenburg Regional Clinical Hospital Named After V. I. Voynov, Orenburg, Russia

Petrozavodsk State University, Petrozavodsk, Russia

Saratov State Medical University named after V.I. Razumovsky, Saratov, Russia

Bryansk Regional Cardiology Clinic, Bryansk, Russia

Kirov Regional Clinical Hospital, Kirov, Russia

Astrakhan Regional Cardiology Dispensary, Astrakhan, Russia

Cardiology Research Center 'Medica', Saint-Petersburg, Russia

Municipal Emergency Hospital, Rostov-on-Don, Russia

‘Family Doctor’, Saint-Petersburg, Russia

Irkutsk State Medical University, Irkutsk, Russia

Moscow Healthcare Department, Municipal Clinical Hospital #13, Moscow, Russia

**Supplementary Table 3.** The *MYBPC3* (NM_000256.3) Variants Identified in Controls

| **Variant** | **Type** | **Novel*** | **Pathogenicity**** | **Count** |
| --- | --- | --- | --- | --- |
| **Pediatrics** | | | | |
| *c.557C>T, p.(P186L)* | Missense | N | VUS | 1 |
| *c.743_746del, p.(D248Afs*51)* | Frameshift deletion | N | P | 3 |
| *c.772+1G>A* | Splice Site | N | P | 1 |
| *c.772G>A, p.(E258K)* | Missense | N | P | 2 |
| *c.1227-13G>A* | Intronic | N | P | 3 |
| *c.1223G>A, p.(S408N)* | Missense | Y | VUS | 1 |
| *c.1273C>T, p.(Q425*)* | Nonsense | N | P | 1 |
| *c.1351+2T>C* | Splice Site | N | P | 3 |
| *c.1482_1483del, p.(R495Gfs*35)* | Frameshift deletion | Y | LP | 1 |
| *c.1483C>T, p.(R495W)* | Missense | N | P | 3 |
| *c.1484G>A, p.(R495Q)* | Missense | N | P | 2 |
| *c.1505G>A, p.(R502Q)* | Missense | N | P | 3 |
| *c.1543_1545del, p.(N515del)* | Inframe deletion | N | LP | 1 |
| *c.1595dup, p.(Q533Pfs*5)* | Nonsense | Y | P | 1 |
| *c.1685C>A, p.(A562E)* | Missense | Y | LP | 1 |
| *c.1786G>A, p.(G596R)* | Missense | N | LP | 1 |
| *c.1855G>T, p.(E619*)* | Nonsense | Y | P | 1 |
| *c.2068-1G>A* | Splice Site | Y | P | 1 |
| *c.2308G>A, p.(D770N)* | Missense | N | P | 1 |
| *c.2373dup, p.(W792Vfs*41)* | Frameshift duplication | N | P | 1 |
| *c.2610del, p.(S871Afs*8)* | Frameshift deletion | N | P | 1 |
| *c.2737+2T>A* | Splice Site | N | P | 1 |
| *c.2827C>T, p.(R943*)* | Nonsense | N | P | 1 |
| *c.2882C>T, p.(P961L)* | Missense | N | VUS | 1 |
| *c.2905+5G>A* | Intronic | N | P | 1 |
| *c.2906-2A>C* | Splice Site | Y | LP | 1 |
| *c.2965del, p.(E989Sfs*3)* | Frameshift deletion | Y | LP | 1 |
| *c.3467dup, p.(P1157Afs*12)* | Frameshift duplication | Y | P | 1 |
| *c.3773T>A, p.(L1258*)* | Nonsense | Y | P | 1 |
| *c.3811C>T, p.(R1271*)* | Nonsense | N | P | 1 |
| **Adults** | | | | |
| *c.334G>T, p.(E112*)* | Nonsense | Y | LP | 1 |
| *c.743_746del, p.(D248Afs*51)* | Frameshift | N | P | 1 |
| *c.772G>A, p.(E258K)* | Missense | N | P | 1 |
| *c.821+1G>A* | Splice Site | N | P | 1 |
| *c.906-36G>A* | Intron | N | P | 1 |
| *c.927-9G>A* | Intron | N | P | 2 |
| *c.966G>A, p.(W322*)* | Nonsense | N | P | 1 |
| *c.971del, p.(I324Tfs*26)* | Frameshift deletion | Y | LP | 1 |
| *c.1037G>A, p.(R346H)* | Missense | N | LP | 6 |
| *c.1120C>T, p.(Q374*)* | Nonsense | N | P | 1 |
| *c.1273C>T, p.(Q425*)* | Nonsense | N | P | 1 |
| *c.1351+2T>C* | Splice site | N | P | 1 |
| *c.1685C>A, p.(A562E)* | Missense | Y | LP | 1 |
| *c.1731G>A, p.(W577*)* | Nonsense | N | P | 2 |
| *c.1790G>A, p.(R597Q)* | Missense | N | P | 1 |
| *c.2345A>G, p.(N782S)* | Missense | Y | VUS | 1 |
| *c.2429G>A, p.(R810H)* | Missense | N | P | 1 |
| *c.2441_2443del, p.(K814del)* | Inframe deletion | N | P | 1 |
| *c.2530_2531del, p.(M844Afs*39)* | Frameshift deletion | N | P | 1 |
| *c.2623C>T, p.(H875Y)* | Missense | Y | VUS | 1 |
| *c.2738-1G>A* | Splice site | Y | P | 1 |
| *c.2778_2781dup, p.(S928Hfs*124)* | Frameshift insertion | Y | P | 2 |
| *c.2827C>T, p.(R943*)* | Nonsense | N | P | 2 |
| *c.2905+1G>A* | Splice site | N | P | 3 |
| *c.2965del, p.(E989Sfs*3)* | Frameshift deletion | Y | LP | 1 |
| *c.3407_3409del, p.(Y1136del)* | Inframe deletion | N | P | 1 |
| *c.3412del, p.(R1138Afs*51)* | Frameshift deletion | Y | P | 1 |
| *c.3763G>A, p.(A1255T)* | Missense | N | P | 1 |
| *c.3773T>A, p.(L1258*)* | Nonsense | Y | P | 1 |
| *c.3790T>C, p.(C1264R)* | Missense | N | LP | 1 |
| *c.3794A>T, p.(E1265V)* | Missense | N | LP | 1 |
| *c.3811C>T, p.(R1271*)* | Nonsense | N | P | 1 |

N – none; Y – yes; P – pathogenic; LP – likely pathogenic; VUS – variant of uncertain significance

*Previously unreported

**Assessed in accordance with the 2015 joint consensus recommendations of ACMG/AMP

**Supplementary Table 4.** Variants in *DES* gene identified in Russian HCM patients

| **Variant** | **Type** | **Novel*** | **Pathogenicity**** | **Count** |
| --- | --- | --- | --- | --- |
| *c.35C>T, p.(S12F)* | Missense | N | P | 1 |
| *c.187G>T, p.(A63S)* | Missense | Y | VUS | 1 |
| *c.206T>C, p.(L69P)* | Missense | N | VUS | 1 |
| *c.216C>A, p.(S72E)* | Missense | Y | VUS | 1 |
| *c.250G>A, p.(G84S)* | Missense | N | VUS | 4 |
| *c.404C>T, p.(A135V)* | Missense | N | VUS | 1 |
| *c.662C>T, p.(A221V)* | Missense | Y | VUS | 2 |
| *c.665G>A, p.(R222H)* | Missense | Y | VUS | 4 |
| *c.833G>A, p.(R278Q)* | Missense | Y | LP | 1 |
| *c.958G>A, p.(E320K)* | Missense | Y | VUS | 1 |
| *c.1049G>A, p.(R350Q)* | Missense | N | P | 1 |
| *c.1063C>T, p.(R355*)* | Nonsense | N | P | 2 |
| *c.1216C>T, p.(R406W)* | Missense | N | P | 1 |
| *c.1243C>T, p.(R415W)* | Missense | N | LP | 1 |
| *c.1256C>T, p.(P419L)* | Missense | N | LP | 1 |
| *c.1286G>A, p.(R429Q)* | Missense | N | VUS | 1 |
| *c.1360C>T, p.(R454W)* | Missense | N | P | 1 |
| *c.1371+1G>A* | Splice Site | Y | LP | 2 |

N – none; Y – yes; P – pathogenic; LP – likely pathogenic; VUS – variant of uncertain significance

*Previously unreported

**Assessed in accordance with the 2015 joint consensus recommendations of ACMG/AMP

**Supplementary Table 5.** Baseline Characteristics of Patients with *MYBPC3*-Associated HCM with Respect to Age

| **Parameter** | ***c.3697C>T* variant** | | | **p-value** | **Other *MYBPC3* variants** | | | | **p-value** | **p-value** | **p-value** |
| --- | --- | --- | --- | --- | --- | --- | --- | --- | --- | --- | --- |
|  | **Overall**  **n = 58** | **Pediatrics**  **n = 15** | **Adults**  **n = 43** |  | **Overall**  **n = 85** | **Pediatrics**  **n=42** | | **Adults**  **n = 43** |  |  |  |
|  | **1** | **2** | **3** | **2-3** | **4** | **5** | | **6** | **1-4** | **2-5** | **3-6** |
| **Demography** | | | | | | | | | | | |
| **Male, n (%)** | 27 (47) | 11 (73) | 16 (37) | **0.019** | 48 (56) | 27 (64) | | 21 (49) | 0.32 | 0.75 | 0.38 |
| **Proband, n (%)** | 53 (91) | 15 (100) | 38 (88) | 0.31 | 79 (93) | 42 (100) | | 37 (86) | 0.98 | 1 | 1 |
| **Age at enrollment, years** | 38 [19;50] | 11.3 [4.8;15.5] | 45 [36;55] | **<0.0001** | 22 [8;47] | 8.1 [2.4;15.2] | | 47 [40;61] | 0.06 | 0.39 | 0.14 |
| **Age at diagnosis, years** | 33 [16;43] | 10.0 [2.0;13.3] | 37 [30;48] | **<0.0001** | 17  [7;44] | 5.5 [0.4;13.0] | | 44 [34;51] | 0.06 | 0.46 | 0.20 |
| **Medical history** | | | | | | | | | | | |
| **Asymptomatic, n (%)** | 10 (17) | 2 (13) | 8 (19) | 1 | 20 (24) | 8 (19) | | 12 (28) | 0.49 | 1 | 0.44 |
| **NYHA class III/IV, n (%)** | 7 (12) | 0 | 7 (17) | 0.17 | 6 (7) | 0 | | 6 (14) | 0.45 | 1 | 0.96 |
| **NT-proBNP, pg/mL** | 509 [140;1369] | 364 [117;1560] | 520 [280;954] | 0.59 | 665  [180;1780] | 425  [185;1031] | | 941 [217;2000] | 0.44 | 0.54 | 0.39 |
| **Arterial hypertension, n (%)** | 17 (29) | 0 | 17 (40) | **0.002** | 19 (22) | 0 | | 19 (44) | 0.46 | 1 | 0.83 |
| **Atrial fibrillation, n (%)** | 8 (14) | 0 | 8 (19) | 0.09 | 11 (13) | 0 | | 11 (26) | 1 | 1 | 0.60 |
| **Stroke in the history, n (%)** | 2 (4) | 0 | 2 (5) | 1 | 1 (1) | 0 | | 1 (2) | 0.56 | 1 | 0.61 |
| **Family cardiomyopathy*, n (%)** | 29 (49) | 8 (53) | 18 (47) | 0.93 | 40 (49) | 23 (55) | | 17 (44) | 1 | 1 | 0.92 |
| **Family SCD*, n (%)** | 9 (17) | 1 (7) | 8 (22) | 0.25 | 10 (12) | 3 (7) | | 7 (18) | 0.58 | 1 | 0.91 |
| **Echocardiography** | | | | | | | | | | | |
| **Asymmetric septal LVH, n (%)** | 46 (81) | 13 (87) | 33 (79) | 0.71 | 59 (69) | 27 (64) | | 32 (74) | 0.19 | 0.18 | 0.85 |
| **LV non-compaction / hypertrabeculation, n (%)** | 6 (11) | 0 | 6 (14) | 0.32 | 7 (8) | 0 | | 7 (16) | 0.86 | 1 | 1 |
| **LVOTO**, n (%)** | 16 (28) | 0 | 16 (38) | **0.006** | 31 (36) | 12 (29) | | 19 (44) | 0.39 | **0.024** | 0.73 |
| **Interventricular obstruction, n (%)** | 3 (5) | 0 | 3 (7) | 0.56 | 10 (12) | 4 (10) | | 6 (14) | 0.24 | 0.56 | 0.48 |
| **Maximal LV wall thickness, mm** | 21±6 | 17±5 | 22±5 | **0.007** | 19±6 | 17±8 | | 21±3 | 0.13 | 0.96 | 0.32 |
| **Extreme LVH ≥ 30mm, n (%)** | 2 (4) | 0 | 2 (5) | 1 | 5 (6) | 5 (12) | | 0 | 0.70 | 0.31 | 0.24 |
| **Restrictive diastole, n (%)** | 7 (13) | 0 | 7 (18) | 0.17 | 6 (7) | 0 | | 6 (15) | 0.43 | 1 | 0.96 |
| **LA dilation, n (%)** | 42 (76) | 9 (60) | 33 (83) | 0.16 | 40 (47) | 9 (21) | | 31 (72) | **0.001** | **0.014** | 0.39 |
| **LV ejection fraction, %** | 69 [63;74] | 74 [72;82] | 66 [60;69] | **<0.0001** | 70 [64;78] | 77 [70;81] | | 65 [60;70] | 0.14 | 0.89 | 0.94 |
| **Hypokinetic HCM***, n (%)** | 5 (9) | 0 | 5 (12) | 0.31 | 1 (1) | 0 | | 1 (2) | **0.038** | 1 | 0.11 |
| **Right ventricular hypertrophy, n (%)** | 9 (17) | 2 (13) | 7 (18) | 1 | 20 (24) | 11 (26) | | 9 (21) | 0.45 | 0.48 | 0.95 |
| **PSAP ≥ 35 mmHg, n (%)** | 15 (28) | 0 | 15 (39) | **0.005** | 13 (15) | 0 | | 13 (30) | 0.10 | 1 | 0.52 |
| **Severe mitral regurgitation, n (%)** | 5 (9) | 0 | 5 (12) | 0.31 | 6 (7) | 0 | | 6 (14) | 0.96 | 1 | 1 |
| **Contrast CMR imaging** | | | | | | | | | | | |
| **Performed, n (%)** | 25 (43) | 6 (40) | 19 (44) | 1 | 47 (55) | 24 (57) | | 23 (53) | 0.21 | 0.40 | 0.52 |
| **LGE on CMR, n (%)** | 20 (80) | 2 (33) | 18 (95) | **0.005** | 27 (58) | 7 (29) | | 20 (87) | 0.10 | 1 | 0.61 |
| **Elecrocardiogram / 24-Hour Holter monitoring** | | | | | | | | | | | |
| **Conduction disturbance****, n (%)** | 15 (27) | 2 (13) | 13 (32) | 0.31 | 27 (33) | | 20 (48) | 7 (17) | 0.59 | **0.029** | 0.20 |
| **WPW, n (%)** | 2 (3) | 2 (13) | 0 | 0.06 | 0 | | 13 (31) | 0 | 0.16 | 0.31 | 1 |
| **PSVT, n (%)** | 25 (45) | 0 | 25 (61) | **<0.0001** | 20 (25) | | 1 (2) | 19 (50) | **0.027** | 1 | 0.45 |
| **NSVT, n (%)** | 17 (29) | 1 (7) | 16 (37) | 0.06 | 20 (24) | | 1 (2) | 19 (50) | 0.56 | 0.46 | 0.66 |
| **Prolonged QTc, n (%)** | 4 (7) | 2(13) | 2 (5) | 0.27 | 12 (14) | | 8 (19) | 4 (9) | 0.76 | 1 | 0.67 |
| **Pharmacological therapy** | | | | | | | | | | | |
| **Beta-blockers, n (%)** | 40 (69) | 9 (60) | 31 (72) | 0.58 | 57 (67) | | 30 (71) | 27 (63) | 0.95 | 0.62 | 0.49 |
| **Antiarrhythmics III class, n (%)** | 9 (16) | 1 (7) | 8 (19) | 0.42 | 6 (7) | | 1 (2) | 5 (12) | 0.18 | 0.46 | 0.55 |
| **Diuretics, n (%)** | 19 (33) | 0 | 19 (44) | **0.001** | 16 (19) | | 0 | 16 (37) | 0.09 | 1 | 0.66 |

NYHA – New York Heart Association; NT-proBNP—N-terminal pro brain natriuretic peptide; SCD – sudden cardiac death; LVH – left ventricular hypertrophy; LV – left ventricle; LVOTO – left ventricular outflow tract obstruction; LA – left atrium; HCM – hypertrophic cardiomyopathy; PASP – pulmonary artery systolic pressure; CMR – cardiac magnetic resonance; LGE – late gadolinium enhancement; PSVT – paroxysmal supraventricular tachycardia; NSVT – no sustained ventricular tachycardia. *in probands only; **both at rest and latent; ***left ventricular ejection fraction < 50%; ****sinoatrial, atrioventricular, and complete bundle branch blocks

**Supplementary Table 6.** Interventions and Outcomes in Patients with *MYBPC3*-Associated Hypertrophic Cardiomyopathy with Respect to Age

| **Parameter** | ***c.3697C>T* variant** | | | **p-value** | **Other *MYBPC3* variants** | | | **p-value** | **p-value** | **p-value** |
| --- | --- | --- | --- | --- | --- | --- | --- | --- | --- | --- |
|  | **All**  ***n* = 58** | **Adults**  ***n* = 43** | **Pediatrics**  ***n* = 15** |  | **All**  ***n* = 85** | **Adults**  ***n* = 43** | **Pediatrics**  ***n* = 42** |  |  |  |
|  | **1** | **2** | **3** | **2 vs 3** | **4** | **5** | **6** | **1 vs 4** | **2 vs 5** | **3 vs 6** |
| **Interventions*** | | | | | | | | | | |
| **Myectomy, n (%)** | 8 (14) | 8 (19) | 0 | 0.10 | 10 (12) | 8 (19) | 2 (5) | 0.92 | 1 | 1 |
| **ICD, n (%)** | 6 (10) | 6 (14) | 0 | 0.32 | 9 (11) | 8 (19) | 1 (2) | 1 | 0.77 | 1 |
| **Appropriate shocks, n (%)** | 2 (3) | 2 (5) | 0 | 1 | 2 (2) | 2 (5) | 0 | 1 | 1 | 1 |
| **Pacemaker, n (%)** | 2 (3) | 2 (5) | 0 | 1 | 1 (1) | 1 (2) | 0 | 0.57 | 1 | 1 |
| **Outcomes** | | | | | | | | | | |
| **Follow-up data available** | 45 (78) | 32 (74) | 13 (87) | - | 82 (96) | 42 (98) | 40 (95) | **-** | **-** | - |
| **Duration of follow-up, years** | 2.3  [0.9-6.1] | 2.2  [0.7-5.5] | 3.3  [1.3-5.8] | - | 3.3  [1.5-5.8] | 4.7  [1.9-7.2] | 3.2  [1.6-5.6] | **-** | **-** | - |
| **All-cause death, n (%)** | 2 (4) | 2 (6) | 0 | 1 | 6 (7) | 4 (10) | 2 (5) | 0.71 | 0.69 | 1 |
| **SCD, n (%)** | 0 | 0 | 0 | 1 | 2 (2) | 0 | 2 (5) | 0.54 | 1 | 1 |
| **Heart transplantation**  **n (%)** | 1 (2) | 1 (3) | 0 | 1 | 2 (2) | 1 (2) | 1 (3) | 1 | 1 | 1 |
| **Hospitalization due to HF**  **n (%)** | 4 (9) | 4 (13) | 0 | 0.31 | 8 (10) | 8 (19) | 0 | 1 | 0.54 | 1 |
| **New-onset stroke, n (%)** | 2 (4) | 2 (6) | 0 | 1 | 3 (4) | 3 (7) | 0 | 1 | 1 | 1 |
| **Composite outcome****  **n (%)** | 6 (13) | 6 (19) | 0 | 0.16 | 14 (17) | 9 (21) | 5 (13) | 0.77 | 1 | 0.32 |

ICD – implantable cardioverter-defibrillator; SCD – sudden cardiac death; HF – heart failure. * Assessed over a single time period, including past history and follow-up; **all-cause death + heart transplantation + new-onset stroke + hospitalization for heart failure.
